# EVALUATING THE IMPACT OF COVID-19 ON THE PREGNANCY AND POSTNATAL PERIOD FOR UK WOMEN

**DOI:** 10.1101/2024.04.29.24306539

**Authors:** Gareth A Nye, George Abou Deb, Seóna Dunne

## Abstract

**INTRODUCTION:** Pregnancy is a crucial period which ultimately directly impacts two individuals health and wellbeing. Within the UK, a standardised pattern of care is established with collaborations across disciplines to the benefit of women and babies. During the COVID19 pandemic, this pattern of care was disrupted to align with protective protocols which until now, has not been formally reported.

**METHODS:** A retrospective, mixed methods study of UK based women pregnant between the years 2012 and 2022 inclusive with no known complications was conducted to collate opinions and experiences of pregnancy with and without the impact of COVID19 restrictions. Quantitative results were analysed using the statistical package GraphPad Prism 9.2.0 and presented as mean values +/- standard deviation were appropriate. In addition, we used a phased approach to open ended questions.

**RESULTS:** Our results showed no significant difference in either the number of appointments or the time of first appointment however an increased percentage of women reported the use of private services during the COVID pandemic. There was no change in the number of midwife appointments during the postnatal period during COVID but there was a significant reduction in the number of health visitor appointments. Overall, the COVID pandemic led to women feeling less satisfied with their care both during their pregnancy and postnatally, but they reported that they continued to be listened to and remained feeling in control of their pregnancy.

**DISCUSSION:** Generally, the changes implemented during the COVID pandemic did not impact women’s pregnancy journey substantially although we have no evidence of the long-term impact on child health and development. Clear themes have been established which can be used to further improve services in maternity and there are key elements to focus on for the future of UK maternity services.

## INTRODUCTION

The impact of SARS-COV2 COVID19 has been felt across the world through the alteration of everybody’s day to day lives ^1,2^. This impact ranges from the devastating effects of the infection itself and the associated loss through to the financial burden on our economies and healthcare ^1-5^.

In the United Kingdom (UK), a national lockdown was announced ^6^ and resulted in significant changes to the hospitals and how their services were delivered with many non-essential services paused ^7,8^. As pregnancy is a time sensitive period, it was essential that maternity care was continued through the pandemic, albeit with significant changes to working practises and unprecedented challenges to keep services running. Initially, as the potential effects of COVID19 were of concern, Antenatal (ANC) clinic appointments were often conducted virtually and restrictions on the number of visitors and/or the presence of a birthing partner was seen universally ^9^.

Previous studies focusing on patient anxiety showed 25.8% of respondents no longer attended in-person visits and due to large uncertainty around the birthing process and COVID-19 restrictions had collectively increased their anxiety ^10^. This was in response to multiple reports of indirect effects of the pandemic leading to combined increased risk of maternal mortality and preterm birth ^10^. We also saw reports on increases in maternal deaths through mental health conditions and suicides which were disproportionately seen in lower income settings ^11,12^.

Now that services have returned to normal within the UK, it is time to retrospectively access and address the feelings of women who were pregnant during this period to look at areas of success in practice and areas to highlight moving forward, looking now towards the extended ramifications on our women and children who lived through the pandemic to gauge future healthcare challenges.

This paper aims to explore the impact of the COVID-19 pandemic on maternity services in the UK, including changes to antenatal, intrapartum and postnatal care, the experiences of pregnant women and healthcare professionals, and the implications for the future of maternity care. By examining the challenges and successes of UK maternity services during the pandemic, this paper aims to contribute to a broader understanding of the impact of COVID-19 on healthcare services and the lessons that can be learned for future pandemics and healthcare crises.

### Study Objectives

The overall objective of this study was to collate the experiences of women who have been pregnant over a 10-year period between 2012 and 2022 to understand the potential impact COVID infection and lockdowns had on a women’s perspective of their care.

## METHODS

### Study design

A retrospective study of women pregnant between the years 2012 and 2022 inclusive was conducted anonymously via a survey distributed on Facebook, Twitter, and pregnancy-related peer and professional communities from 20^th^ February-20^th^ March 2022. Survey was designed to contain both quantitative and qualitative data through mixed methods of closed and open questioning and followed the Checklist for Reporting Results of Internet E-Surveys (CHERRIES) in its design and analysis ^13^.

### Setting

English-language online survey was open to any woman based within the United Kingdom at the time of their pregnancy.

### Participants

Self-identified women who had been pregnant during the previous decade and were able to complete an online survey in English, recruited via Facebook and Twitter, including pregnancy-specific Facebook groups. The survey link was also shared with pregnancy-specific professional communities for distribution through their networks of pregnant women. For this paper, any women reporting a complication during their pregnancy (e.g. Gestational diabetes, hypertension or pre-eclampsia), or women which pre-existing conditions which would result in non-standard care during pregnancy were excluded from this report and will form the basis of future publications.

### Demographics and basic questions

Participants were asked to complete questions based on maternal demographics and geographic locations. Additional pregnancy specific questions were asked to cover both the pregnancy and post natal period. General questions related to the COVID pandemic and vaccine were asked to all respondents with additional specific questions around COVID infection and changes to lifestyle during the pandemic asked to only respondents pregnant during the years 2020 to 2022 inclusive. Questions relating to participants feelings of satisfaction or otherwise were presented as a 1-10 numerical scale with 1 being the least and 10 being the most. Questions analysed for this study were as follows - How satisfied were you with the care you’ve received during pregnancy? How in control of your pregnancy did you feel? How do you feel your thoughts and concerns where managed during your pregnancy? How do you feel your postnatal care was?

### Data analysis

Quantitative results were analysed using the statistical package GraphPad Prism 9.2.0 and presented as mean values +/- standard deviation were appropriate. Percentages were converted to the nearest whole number and presented as pre-2020 vs 2020-2022 values. Only uncomplicated pregnancies are reported in this study to exclude additional factors which may influence maternity care.

### Thematic analysis

We used a phased approach to open ended questions inspired by Braun and Clarke^14^ to identify patterns and themes. The phases are referred to; (1) Familiarisation with the data. (2) Generating themes. (3) Reviewing themes. (4) Defining and naming themes. (5) producing the report ^14^. We carried out the process of coding the material in a flexible way and moved back and forth through the phases as necessary ^15^

### Ethics

Ethical approval for this study was granted by the University of Chester Faculty of Medicine and Life Sciences Research Ethics Committee. Participants were asked to read and sign a participant information sheet before completing the survey. Data was anonymous at the point of survey completion.

## RESULTS

### Patient demographics

This survey returned a total of 274 respondents with 124 individual responses in the pre-2020 group and 154 responses pregnant during 2020-2022. There was an increased response rate from the North West England accounting for 40% of all responses with low response rates in Northern Ireland and Scotland. There were no significant difference in the mean maternal age of both groups with an average of 29.8 (+/- 5.0) years and 31.5 (+/- 4.6) years in the pre-2020/2020-2022 groups respectively. There was a higher rate of first-time pregnancies in the pre-2020 group (69% to 61%).

There were no significant differences in either the approximate number of appointments, or the approximate time of the first appointment in either group, but there was raise in the percentage of respondents using private care (11.8% vs 14.6%) and private scans (37% vs 55%) during 2020-2022 when compared with the pre-2020 groups.

Interestingly, an increased percentage of births ending in planned c-sections were seen within the 2020-2022 group but no change in the percentage of midwife led vaginal deliveries. There were no changes in birth location nor any significant changes in mean birth weight.

There were differences in main feeding method at birth with a lower number of mothers choosing to breast fed in the 2020-2022 group (Bottle fed 18% vs 21%, Breast fed 72% vs 63%, Combination 11% vs 15%), however by 6 months of age the percentage of mothers breast feeding in the 2020-2022 group was higher than in the pre-2020 group (Bottle fed 50% vs 43%, Breast fed 37% vs 48%, Combination 11% vs 8%).

### Postnatal care

No significant change in the number of midwife visits were reported between the groups and a higher percentage of women in the 2020-2022 reported having contact with a health visitor (91% vs 96%). However, women saw a decrease in health visitors attending in person appointments (91% vs 77%) and a rise in phone/remote appointments (5% vs 23%) in the 2020-2022 group. A significant decrease in health visitor appointments were reported during this time (3.5 +/- 3.9 vs 2.6 +/- 1.8, P = 0.05).

### Overall Satisfaction

For each of the four questions asked related to the participants perception of their pregnancy, lower average self-reported scores from the group were seen in those pregnant during the years 2020-2022.

Significantly lower scores were reported for patient satisfaction in care during pregnancy, during the COVID time period (8.11 +/- 1.91 vs 7.26 +/- 2.20, P = <0.01) and significantly lower self reported scores for postnatal care during the same period (7.31 +/- 2.69 vs 5.95 +/- 2.94, P = <0.01).

Level of control remained non significant, as did the management of concerns during pregnancy.

### Impact of COVID pandemic on pregnancy period

80% of respondents who were pregnant during the COVID pandemic faced a lockdown during their pregnancy to some extent, with 45% of respondents being under a lockdown during the time of delivery.

60% of those who answered faced a rearrangement of appointments to remove the face to face aspect with an average satisfaction score of 5.8 (+/-2.9) with these appointments, when using a 10 point scale.

Despite only 7% of respondents contracting COVID-19 during their pregnancy, 78% were concerned about the pandemic. 45% saw changes to the original birth plan and 14% of respondents shared that the pandemic influenced their decision making about infant feeding.

### Thematic analysis

#### Utilization of Private Care or scans during Pregnancy (pre-COVID-19)

The thematic analysis unveiled that the majority of participants refrained from seeking private healthcare services during pregnancy. However, a notable subset of individuals opted for private care, driven by a diverse range of motivations. These motivations encompassed a preference for personalized one-to-one care, concerns regarding the accessibility of specific diagnostic tests, a desire for enhanced control over their healthcare decisions, proactive anxiety management, and a need for additional support and reassurance. Particularly, individuals with a history of miscarriage emphasized the significance of these factors. Key determinants for private scans included the desire for gender determination, the acquisition of multimedia keepsakes, and emotional reassurance with accessibility and timing of National Health Service (NHS) scans playing pivotal roles in respondents’ decisions. These findings underscored the intricate and multifaceted nature of the factors that shape individuals’ choices concerning pregnancy care.

#### Pregnancy Care During the COVID-19 Pandemic

In the context of the COVID-19 pandemic, the thematic analysis revealed that the predominant trend among participants during their pregnancy was the avoidance of seeking private healthcare services. Nevertheless, a minority of participants opted for private care, driven by various reasons, including prior miscarriages, pregnancy-related concerns, COVID-19 restrictions, and previous traumatic birth experiences. Some participants sought private care to ensure added assurance or to involve their partners in the pregnancy journey. The primary motivation for seeking private care predominantly stemmed from COVID-19 restrictions, which hindered access to NHS scans. This analysis underscored the multifaceted rationales for seeking private prenatal scans, encompassing reassurance, early gender determination, partner participation, and convenience. Notably, cost emerged as a potential barrier for certain families. The identified themes emphasized the complexity of the labour and delivery process, often necessitating medical interventions to ensure safety. This further underscored the importance of offering individualized and flexible healthcare options during pregnancy to accommodate the unique needs and preferences of expectant parents. It is noteworthy that the adoption of virtual appointments became prevalent during the pandemic, particularly for antenatal check-ups and health visitor assessments. However, some consultations with specialists remained in-person, and a subset of participants engaged in a combination of virtual and in-person appointments, highlighting the evolving landscape of healthcare delivery.

#### Impact of the COVID-19 Pandemic on Birth Plans

The COVID-19 pandemic exerted a substantial influence on the birth plans of many respondents, resulting in a spectrum of modifications. These modifications ranged from restrictions on birth partners and visitors to changes in birthing locations and alterations in antenatal and postnatal care. The emotional and logistical challenges stemming from these changes underscored the importance of adaptability and flexibility during pregnancy and childbirth, particularly in times of uncertainty. The majority of respondents expressed concerns about various issues related to the pandemic, including anxieties surrounding partner restrictions during childbirth, limitations on visitation, and the broader impact of COVID-19 on pregnant women. Additional concerns included maternity ward closures and inadequate access to information. While some respondents were relatively unperturbed by pandemic-related issues, the prevailing sentiment was one of apprehension regarding the potential impact of the pandemic on their pregnancy and birthing experiences.

#### Infant Feeding Decisions during the Pandemic

In terms of infant feeding decisions, the pandemic did not exert a significant influence on most participants. The majority reported a pre-existing determination to breastfeed that remained unaffected by the pandemic. A subset of participants mentioned that the pandemic heightened their resolve to breastfeed, while others perceived a lack of breastfeeding support as a challenge. A minority of participants made the decision to formula feed, and these cases were relatively infrequent. Overall, the pandemic did not substantially impact infant feeding decisions.

#### COVID-19 Infections during Pregnancy

The thematic analysis of COVID-19 experiences during pregnancy revealed that the majority of respondents did not contract the virus, with only a few reporting infections. Hospital admissions due to COVID-19 during pregnancy were rare. Some mentioned losing their sense of taste without a positive test. Infections occurred at various gestational weeks, and most pregnancies proceeded without significant COVID-19-related complications.

#### Reasons for Not Being Fully Vaccinated Against COVID-19

Based on when the participant was pregnant, we see two very clear themes emerging. For those pregnant during the pre-COVID time period, answers included vaccine hesitancy or the reliance on natural immunity as reasons against, with others emphasising the importance of vaccination in safeguarding both personal health and the broader community. Taking responses from participants who were pregnant during the COVID period we see a change in themes with common themes among those who had not completed vaccination including concerns about breastfeeding, conflicting information, hesitancy due to the perceived experimental nature of vaccines during pregnancy, lack of research during pregnancy, and doubts about vaccine efficacy and transmission prevention. Some individuals mentioned waiting until they finished breastfeeding, receiving medical advice from midwives, having a needle phobia, relying on natural antibodies from previous infections, or opting not to disclose their reasons. These themes illuminate the complex factors shaping vaccination decisions during pregnancy.

## DISCUSSION

In this study, a significant decrease in maternal satisfaction with the care received during pregnancy was observed in the period of 2020-2022 compared to pre-2020 levels. This may be attributed to the changes in the delivery of maternity services due to the COVID-19 pandemic. No significant difference in the level of control mothers felt over their pregnancy were reported suggesting that the COVID induced changes to maternity services did not impact women’s sense of control.

There was a trend towards an increase in maternal age in the 2020-2022 period compared to pre-2020, although this was not statistically significant. This may reflect changing social trends, such as women delaying pregnancy for career or personal reasons or as this study observed a decrease in the proportion of first pregnancies in the 2020-2022 period compared to pre-2020, it could be women are reporting second pregnancies during COVID.

No significant differences were reported in the number of appointments attended or the timing of the first appointment between the two time periods studied which shows the ability of our healthcare system to maintain a standard level of care. An increase in the proportion of respondents who reported using private scans in the 2020-2022 period compared to pre-2020 was noted and is strongly suggested to be related to changes in the availability or accessibility of NHS scans during the pandemic. However, it is important to note that the use of private scans was still a minority practice overall.

In terms of mode of delivery, no significant differences were reported in the proportion of emergency or planned C-sections or vaginal deliveries between the two time periods studied. This suggests that changes to maternity services during the COVID-19 pandemic did not significantly impact the method of delivery for most women.

Finally, this study saw a decrease in the proportion of mothers who reported exclusively breastfeeding at birth and at 6 months postpartum in the 2020-2022 period compared to pre-2020. This may be reflective of changes to maternity services during the pandemic, such as decreased access to lactation support or changes in hospital policies related to breastfeeding. It might also be due to worries around the passing of COVID through breast milk from mother to baby. This indicates however a clear issue during the COVID time period and should be focused on to improve for any future pandemics.

### Post-natal care

The postnatal care provided by midwives and health visitors is an important aspect of maternity services. In this study, we examined the changes in postnatal care during the COVID-19 pandemic in the UK. The results showed that there was a slight decrease in the mean number of midwife visits to the home (3.1 +/- 2.0 visits pre-2020 vs 2.7 +/- 1.9 visits in 2020-2022), but this difference was not statistically significant (p=0.23).

In terms of health visitor contacts, there was a significant decrease in the mean number of contacts (3.5 +/- 3.9 pre-2020 vs 2.6 +/- 1.8 in 2020-2022, p=0.05). This decrease could be due to several factors, such as increased workload for health visitors during the pandemic or changes in the way health visitor services were delivered during lockdowns. Interestingly, there was a significant increase in the percentage of respondents who reported having contact with a health visitor during the pandemic compared to pre-2020 (96% in 2020-2022 vs 91% pre-2020, p<0.01) which may have been in response to the move to virtual appointments.

## CONCLUSION

The results of this study suggest that the COVID-19 pandemic has had a negative impact on maternal satisfaction with care during pregnancy and postnatal care. The decrease in satisfaction with care during pregnancy and postnatal care is a cause for concern, as this may impact the physical and mental health outcomes of mothers and infants. The pandemic may have contributed to increased anxiety and stress for pregnant and postnatal women, which may have impacted their perceptions of care.

The reduction in face-to-face consultations may have led to a lack of personal connection with healthcare professionals and decreased opportunities for discussion and support. Virtual consultations may not have been sufficient to replace in-person consultations and may have contributed to a perception of a lack of support. Women may have also found it difficult to access information about available services and may have experienced difficulties accessing appropriate care due to reduced staffing levels and resources.

Additionally, the study found a significant decrease in maternal perception of how their thoughts and concerns were managed during pregnancy, suggesting that healthcare professionals may need to improve their communication and support for women during the pandemic. During the pandemic, it was challenging for healthcare professionals to provide the same level of support as they did pre-pandemic due to increased demands on their time, resources, and a lack of personal contact with patients. Virtual consultations may have also contributed to difficulties in communication, as non-verbal cues and physical examination were not possible.

Despite the challenges posed by the pandemic, there was no significant difference in maternal perception of control during pregnancy, indicating that women still felt in control of their pregnancies. This may be due to the availability of virtual consultations and the continued provision of essential antenatal care. It may also reflect the resilience of women during the pandemic and their ability to adapt to changes in healthcare services.

## Data Availability

All data produced in the present study are available upon reasonable request to the authors

